# Influence of nutritional status on eating habits and food choice determinants among Brazilian women during the COVID-19 pandemic

**DOI:** 10.1101/2020.11.03.20225136

**Authors:** Bruna Caruso Mazzolani, Fabiana Infante Smaira, Gabriel Perri Esteves, Heloísa C. Santo André, Milla Cordeiro Amarante, Daniela Castanho, Karen Campos, Fabiana Braga Benatti, Ana Jéssica Pinto, Hamilton Roschel, Bruno Gualano, Carolina Nicolleti Ferreira

## Abstract

We aimed to evaluate the influence of nutritional status on eating habits and food choice determinants among Brazilian women during the COVID-19 outbreak. This cross-sectional survey was conducted between June and September, 2020, period in which social distancing measures were in place. Participants (n=1,183) were classified as normal weight (60.4%), overweight (26.2%) and obese (13.4%). Eating habits changed during quarantine irrespective of nutritional status. The number of women participating in grocery shopping was reduced by 34% during quarantine (p<0.001, OR=0.55, 0.79), whereas participation in cooking and ordering delivery service increased by 28% (p=0.004, OR=1.08, 1.51) and 146% (p<0.001, OR=2.06, 2.95), respectively. The number of participants reporting the habit of snacking (p=0.005, OR=1.07, 1.43) and eating at the table increased by 24% and 40% (p<0.001, OR=1.20, 1.64). Interestingly, the number of participants reporting the habit of dieting decreased by 41% (p<0.001, OR=0.59 [0.50, 0.70]). During the quarantine, “liking”, “need and hunger”, and “habits” were the most commonly reported determinants of food choice overall. “Health”, “natural concerns” and “need and hunger” were less important determinants for participants with overweight/obesity compared to those with normal weight. Regression models showed that *(i)* “health”, “natural concerns” and “affect regulation”; *(ii)* “health”, “pleasure”, “convenience”, and “natural concerns”; and *(iii)* “visual appeal” and “pleasure” were the food choice determinants more associated with eating habits among women with normal weight, overweight and obesity, respectively. In conclusion, eating habits were influenced during the pandemic despite nutritional status, whereas food choice determinants differed between overweight/obesity and normal weight women.

## 1. INTRODUCTION

Quarantine and social isolation are associated with unpleasant experiences, including distancing of family and friends, loss of freedom, uncertainty about one’s illness, and boredom (Brooks, et al., 2020). To contain the COVID-19 pandemic, social distancing has been imposed worldwide. As a consequence, psychological symptoms, stress, irritability, insomnia, feelings of anger, and emotional exhaustion have been reported (Brooks, et al., 2020). These changes in emotional state can potentially modify lifestyle habits, including eating habits.

It has been suggested that routine changes can have a greater impact in women since they hold more responsibilities regarding family food choices (Herman & Polivy, 2010). In addition, women tend to eat more in stressing situations when compared to men (Bennett, Greene, & Schwartz-Barcott, 2013). A study evaluating choices, behaviors and food preferences among women with normal weight, overweight or obesity showed that: *i)* Taste is fundamental for food choice irrespective of BMI; *ii)* Price was more determinant for food choice among women with overweight and obesity *vs*. normal weight; *iii)* Health was an influential factor in food choice mostly among the participants with normal weight; and *iv)* women with overweight and obesity report liking a wider variety of foods, both healthy and less healthy, and mentioned using food more often as a mechanism to cope with periods of stress, depression or boredom, whereas their normal weight peers reported eating less or the same amount of food when facing negative emotions (Dressler & Smith, 2013). These data collectively suggest that nutritional status plays an important role on food choices and preferences among women.

Eating habits and the complex process of food choices are driven by an interplay of physiological mechanisms, genetics, epigenetics, economic and behavioral factors, and organoleptic characteristics of foods (Higgs & Thomas, 2016; Leng, et al., 2017; Nolan, Halperin, & Geliebter, 2010). The COVID-19 pandemic resulted in profound changes in women’s daily routine which may potentially influence eating habits in unpredictable ways. Whether and to which extent nutritional status are associated with modifications in eating habits during the pandemic remains unknown.

The aim of this study was to evaluate the influence of nutritional status on eating habits and food choice determinants among Brazilian women during the COVID-19 outbreak.

## 2. METHODS

### 2.1 Study design and participants

This is a cross-sectional survey conducted between June and September 2020, period in which a set of social distancing measures to contain the spread of COVID-19 were in place in Brazil. Participants were recruited through advertisements on social medias (Facebook^®^, WhatsApp^®^, Instagram^®^, Twitter^®^), press release, television, journals, and radio. Inclusion criteria were as follows: women aged ≥ 18 years, currently living in Brazil, with ability to read, and with internet access.

All participants completed an online survey on Google^®^ Forms platform (Google^®^ LLC, Menlo Park, CA, USA), which enquired about their demographic, socioeconomic, and anthropometric characteristics, psychological symptoms, lifestyle and eating habits.

### 2.2 Evaluation tool

The online survey included questions categorized into the following sections: *i)* participants characteristics (age, ethnicity, marital status, educational level, smoking, and chronic medical conditions); *ii)* anthropometric data (self-reported weight and height); *iii)* eating habits before and during the COVID-19 quarantine (cooking, participation in grocery shopping, use of delivery services, alcohol consumption, habits of snacking, replacing main meals for snacks, eating at table, eating in front of television/tablet/cellphone, and dieting).

Subsequently, participants filled the following inventories: *i)* the Brazilian Portuguese version of The Eating Motivation Survey (TEMS) (Sproesser, Moraes, Renner, & Alvarenga, 2019) to evaluate determinants of food choices; *ii)* the Binge Eating Scale (BES) to evaluate symptoms of binge eating episodes (Silvia Freitasa, 2001); *iii)* the short version of Disordered Eating Attitute Scale to evaluate eating attitudes (Alvarenga, Santos, & Andrade, 2020); *iv)* the Depression Anxiety Stress Scale-21 (DASS-21) to assess depression, anxiety and stress symptoms (Vignola & Tucci, 2014); and *v)* the UCLA Loneliness Scale to assess perceived feeling of loneliness (Barroso, Andrade, Midgett, & Carvalho, 2016).

### 2.3. Data privacy and Ethics aspects

This study was approved by the local ethical committee and was conducted in accordance with the Helsinki declaration. Approved Informed Consent Form was signed digitally by all participants before initiating the survey.

### 2.4 Statistical analysis

Descriptive data are presented as mean and 95% confidence interval (95%CI) for continuous variables and absolute and relative frequency (n[%]) or odds ratio (OR) and 95%CI for categorical variables. Potential changes in eating habits (cooking, participation in grocery shopping, use of delivery services, alcohol consumption, habits of snacking, replacing main meals for snacks, eating at table or in front of television/tablet/cellphone, and dieting) were assessed by generalized estimating equations (GEE) model, based on the assumption of a binomial distribution, logit link function, and an exchangeable working correlation, with time (before and during social distancing) as fixed factor and subjects as random factor. Potential changes in determinants of food choices, binge eating symptoms, eating attitudes, and psychological symptoms were assessed by generalized estimating equations (GEE) model, based on the assumption of a normal distribution, identity link function, and an exchangeable working correlation, with group (normal weight, overweight and obese) as fixed factor. All GEE models were adjusted for age, educational level, ethnicity, marital status, and number of comorbidities. Tukey’s post-hoc test was used for multiple-comparison correction. Finally, associations between eating habits (independent variables) and determinants of food choices (dependent variables) were tested using linear regression models. All regression models were adjusted for age, BMI, educational level, ethnicity, marital status, and number of comorbidities. All analyses were performed using the statistical package SAS (version 9.4). The level of significance was set at p≤0.05.

## 3. RESULTS

One thousand two hundred and forty-one participants completed the online survey; 1,183 participants had valid data and were included in the analysis. Age ranged between 18 and 72 years (34.56 [33.85, 35.27]). Most participants were white (77.8%), single (55.5%) and had higher educational level (72.4%). According to BMI, 60.4%, 26.2% and 13.4% were classified as normal weight, overweight and obese, respectively. Age (p<0.001), ethnicity (p=0.012), marital status (p<0.001), educational level (p=0.007), and self-reported prevalence of comorbidities (all p< 0.050) were different between groups (Supplementary Table 1).

Differences in eating habits between BMIs were not influenced by quarantine. However, when data were polled irrespective of nutritional status, the number of women participating in grocery shopping reduced by 34% (p<0.001, OR=0.66 [0.55, 0.79], Figure 1) during social distancing, whereas participation in cooking and ordering delivery service increased by 28% (p=0.004, OR=1.28 [1.08, 1.51], Figure 1) and 146% (p<0.001, OR=2.46 [2.06, 2.95], Figure 1), respectively. The number of participants eating at the table increased by 40% (p<0.001, OR=1.40 [1.20, 1.64], Figure 1), but no changes where observed for eating in front of TV/tablet/cellphone (p=0.693, Figure 1). No changes were observed for the habit of replacing main meals for snacks (p>0.050, Figure 1A), but there was an increase in the number of participants reporting the habit of snacking (p=0.005, OR=1.24 [1.07, 1.43], Figure 1). Interestingly, however, the number of participants reporting the habit of dieting decreased by 41% (p<0.001, OR=0.59 [0.50, 0.70], Figure 1). Also, the number of participants consuming alcohol beverages reduced during social distancing (p < 0.001, OR = 0.56 [0.49, 0.63], Figure 1).

**Figure 1.**
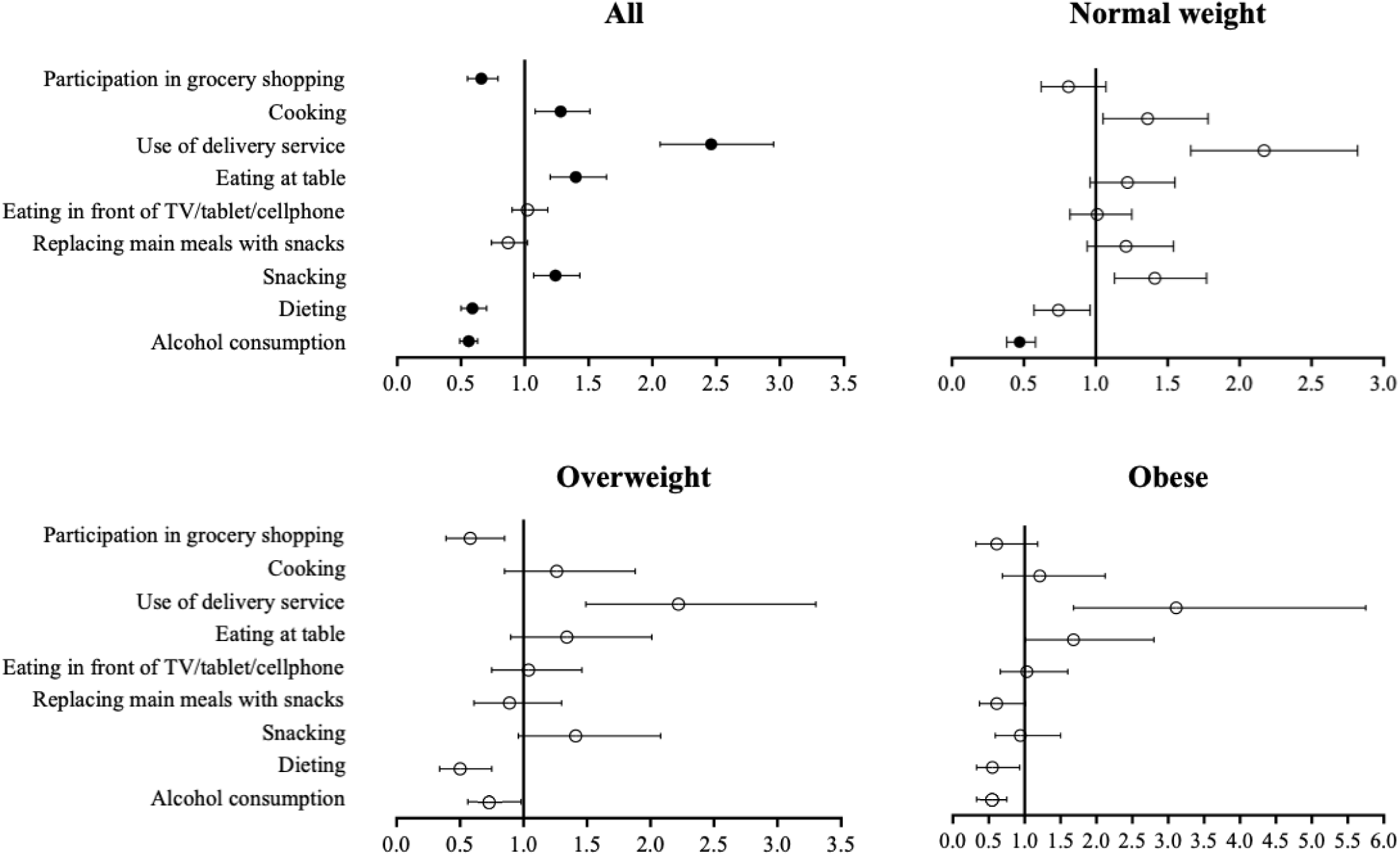
Changes in eating habits during the COVID-19 pandemic. Data are presented as odds ratio (OR) and 95%CI; ○: p>0.05; •: p<0.05 for main effect of time (before to during pandemic). There were no significant group*time interactions.

Before quarantine, the number of overweight and obese women that reported engaging in restrictive diets was 102% (p<0.001) and 129% (p<0.001) higher than normal weight group, respectively. Similarly, replacing meals was 109% (p<0.001) and 208% (p<0.001) higher in both groups when compared to normal weight group. During quarantine, overweight group remained replacing meals more frequently (55%) than normal weight group (p=0.04).

“Liking”, “need and hunger”, and “habits” were the most commonly reported determinants of food choices for all groups. “Health”, “natural concerns” and “need and hunger” were less important determinants for participants with overweight and obesity compared to those with normal weight (all p<0.001, Table 1). Additionally, “habits” were less important for participants with obesity compared to those with normal weight (p<0.001, Table 1). In contrast, “affect regulation” was a more important determinant for participants with overweight and obesity compared to those with normal weight (both p<0.001), “Health”, “natural concerns” and “affect regulation” were less important for participants with overweight than obesity (all p<0.050, Table 1). Disordered eating attitudes and symptoms of binge eating episodes, depression and anxiety were more prevalent in participants with overweight and obesity, compared to those with normal weight (all p<0.050, Table 1). Moreover, all of these symptoms were more prevalent in participants with obesity than in those with overweight (all p<0.050, Table 1). Stress was more prevalent among participants with obesity compared to those with normal weight (p<0.001, Table 1).

**Table 1.** Food choice determinants and psychological symptoms.

|  | Normal weight | Overweight | Obese |
| --- | --- | --- | --- |
| *TEMS (I eat what I eat ...)* |  |  |  |
| Liking | 3.00 (2.95, 3.05) | 2.92 (2.84, 3.00) | 2.93 (2.82, 3.04) |
| Health | 2.55 (2.48, 2.61) | 2.25 (2.15, 2.35)<sup>a</sup> | 1.96 (1.82, 2.10)<sup>a,b</sup> |
| Natural concerns | 1.95 (1.88, 2.02) | 1.79 (1.68, 1.90)<sup>a</sup> | 1.50 (1.35, 1.65)<sup>a,b</sup> |
| Need and Hunger | 2.81 (2.75, 2.86) | 2.53 (2.45, 2.61)<sup>a</sup> | 2.38 (2.26, 2.49)<sup>a</sup> |
| Habits | 2.75 (2.69, 2.81) | 2.58 (2.49, 2.67) | 2.43 (2.30, 2.55)<sup>a</sup> |
| Pleasure | 2.02 (1.96, 2.08) | 2.13 (2.04, 2.23) | 2.18 (2.05, 2.32) |
| Convenience | 1.95 (1.89, 2.00) | 1.88 (1.78, 1.98) | 1.93 (1.80, 2.05) |
| Weight control | 1.46 (1.39, 1.52) | 1.56 (1.46, 1.65) | 1.41 (1.29, 1.55) |
| Sociability | 1.65 (1.58, 1.72) | 1.72 (1.61, 1.82) | 1.57 (1.42, 1.71) |
| Traditional Eating | 1.73 (1.67, 1.80) | 1.74 (1.65, 1.84) | 1.71 (1.57, 1.84) |
| Price | 1.49 (1.42, 1.56) | 1.37 (1.27, 1.47) | 1.50 (1.36, 1.64) |
| Visual Appeal | 1.08 (1.02, 1.14) | 1.18 (1.09, 1.27) | 1.20 (1.07, 1.32) |
| Affect Regulation | 1.01 (0.94, 1.08) | 1.30 (1.20, 1.41)<sup>a</sup> | 1.65 (1.51, 1.80)<sup>a,b</sup> |
| Social Norms | 1.09 (1.04, 1.15) | 1.04 (0.95, 1.13) | 1.18 (1.06, 1.30) |
| Social Image | 0.41 (0.36, 0.45) | 0.46 (0.40, 0.53) | 0.48 (0.39, 0.57) |
| *DEAS score* | 27.55 (26.81, 28.29) | 31.36 (30.25, 32.48)<sup>a</sup> | 35.28 (33.71, 36.82)<sup>a,b</sup> |
| *BES score* | 7.01 (6.49, 7.52) | 11.51 (10.73, 12.30)<sup>a</sup> | 15.41 (14.32, 16.51)<sup>a,b</sup> |
| *Depression symptoms score* | 5.72 (5.32, 6.12) | 6.16 (5.55, 6.76)<sup>a</sup> | 7.30 (6.46, 8.14)<sup>a</sup> |
| *Anxiety symptoms score* | 3.82 (3.49, 4.16) | 4.32 (3.81, 4.83)<sup>a</sup> | 5.73 (5.02, 6.45)<sup>a,b</sup> |
| *Stress symptoms score* | 7.74 (7.34, 8.13) | 8.28 (7.68, 8.88)<sup>a</sup> | 9.30 (8.47, 10.14)<sup>a</sup> |
| *Loneliness Scale score* | 30.81 (29.88, 31.74) | 31.38 (29.97, 32.79) | 34.11 (32.14, 36.09)<sup>a</sup> |

Data presented as mean and 95% confidence interval (95%CI). <sup>a</sup> $p < 0.05$ vs. normal weight; <sup>b</sup> $p < 0.05$ vs. overweight. Abbreviations: DEAS, Disordered Eating Attitude Scale; BES, Binge Eating Scale. Note: For The Eating Motivation Survey (TEMS), the sub-items for assessing motives for eating behavior were introduced by the following item stem 'I eat because ...' or by 'I select certain foods because ...' and answers were given on a seven-point rating scale from 1 'never' to 7 'always'.

Linear regression models showed that “health”, “natural concerns” and “affect regulation” were the food choice determinants more associated with eating habits (≥ 3) among women with normal weight (Figure 2). “Health”, “pleasure”, “convenience”, and “natural concerns” were the determinants more associated with eating habits among women with overweight, whereas “visual appeal” and “pleasure” were those more associated with eating habits among participants with obesity (Figure 2).

**Figure 2.**
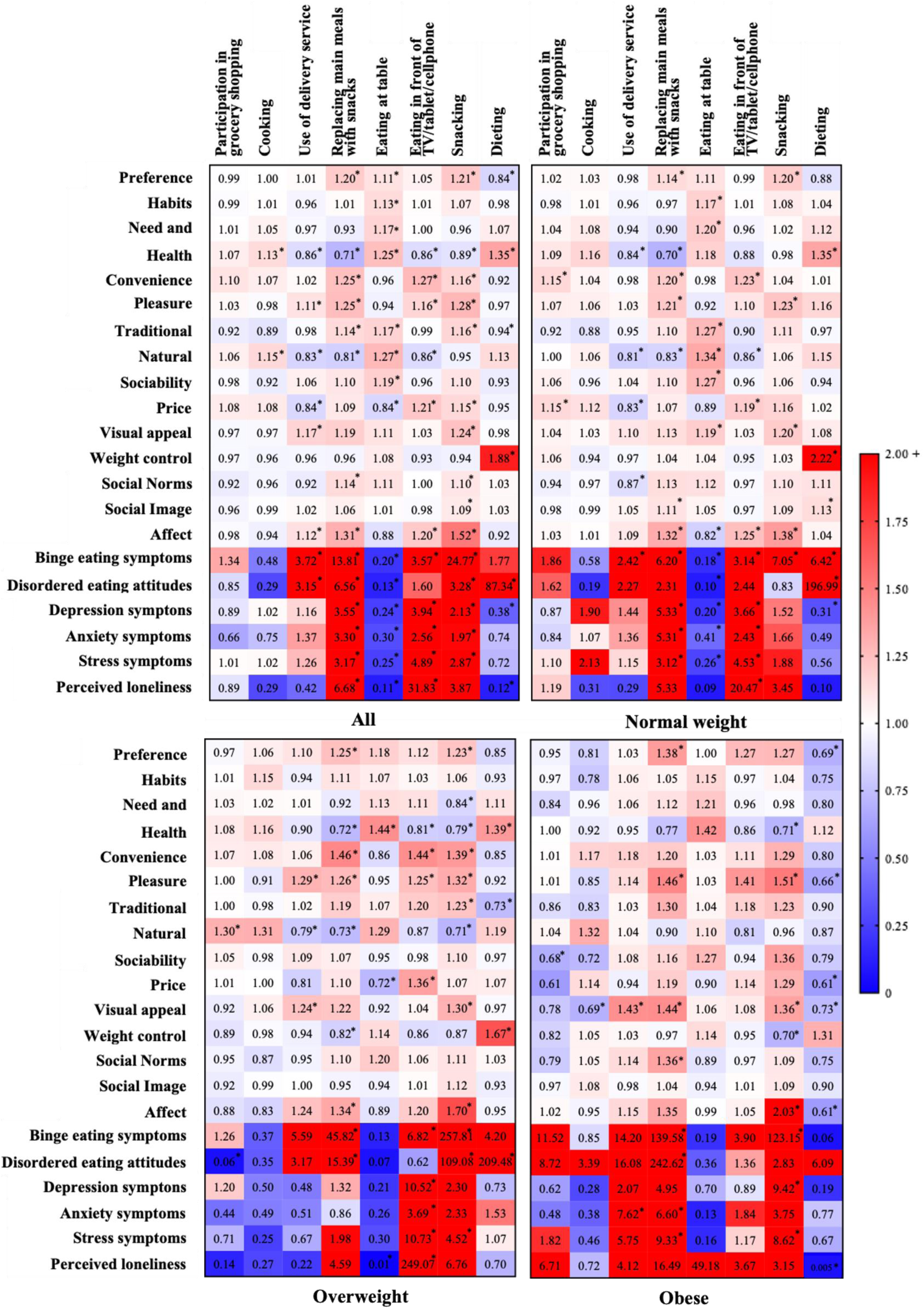
Associations between eating habits, food choice determinants and psychological symptoms. Data are presented as odds ratio (OR). *p<0.05.

Interestingly, “weight control” was not associated with any eating habits, with exception of dieting in women with normal weight (p<0.001, OR= 2.22 [1.89, 2.59], Figure 2) and those with overweight (p<0.001, OR=1.67 [1.37, 2.04], Figure 2). Psychological symptoms were associated with worse eating habits (e.g., replacing main meals, eating in front of TV/tablet/cellphone, and snacking) in all groups. Of relevance, disordered eating attitudes were strongly associated with dieting among participants with normal weight (p<0.001, OR= 196.99 [35.88, 1081.37], Figure 2) and overweight (p<0.001, OR= 209.48 [12.71, 3452.66], Figure 2). Overall, associations were similar between groups in terms of magnitude and direction, although some of them did not reach statistical significance in overweight and obesity groups, which had a smaller sample size.

## 4. DISCUSSION

The main findings of this study were that: *i)* eating habits changed among women irrespective of nutritional status during quarantine due to COVID-19 in Brazil; however, *ii)* the determinants of food choice differed as a function of nutritional status during quarantine. Furthermore, *iii)* the determinants of food choice and psychological symptoms were associated with eating habits, with some associations being affected by nutritional status.

The COVID-19 pandemic and the set of social distancing measures adopted around the world have resulted in dramatic changes in daily living and lifestyle. A growing number of studies have shown that diet has also changed during this period in several countries (Ammar, et al., 2020; Mitchell, Yang, Behr, Deluca, & Schaffer, 2020; Pellegrini, et al., 2020). This study extends this notion to show that, although eating habits did not appear to be influenced by nutritional status, food choice determinants considerably differed across nutritional status categories.

It has been shown that food choices and preferences are differently affected in women with obesity and overweight as compared with those with normal weight (Dressler & Smith, 2013). As women with overweight and obesity commonly use food more often as a mechanism to cope with periods of stress, depression or boredom (Dressler & Smith, 2013), one could expect that nutritional status would be a factor influencing eating habits changes during the COVID-19 pandemic. During the pandemic, women participating in grocery shopping reduced by 34%, whereas participation in cooking and ordering delivery service increased by 28% and 146%, respectively. The number of participants reporting the habit of snacking and eating at table increased by 24% and 40%, respectively, while the number of participants reporting the habit of dieting decreased by 41%. These latter changes could be an attempt to cope with the emotional stress related to the social isolation due to the pandemic. Importantly, in contrast to our hypothesis, the adaptations in eating habits were comparable among women with normal weight, overweight and obesity.

Conversely, we observed that food choice determinants potentially able to affect eating habits were differently reported between women with obesity/overweight and those with normal weight. For instance, women with obesity/overweight, as compared with their peers with normal weight, chose foods preferentially to deal with their emotions (*e.g*., “affect regulation”) rather than guided by physiological signs (*e.g*., “need and hunger” and “health concerns”). However, these food choice determinants in isolation may have not been sufficient to modify eating habits. It is important to bear in mind that eating habits are driven by a complex, multifactorial process that involves a variety of behavioral and biological aspects (Higgs & Thomas, 2016; Leng, et al., 2017; Nolan, et al., 2010). In this study, food choice determinants associated with social and economic factors (*e.g*., “price”, “convenience”, “sociability”, “social norms”, “social image”) did not differ as a function of nutritional status. This could partially explain the absence of major differences in eating habits during the pandemic between women with obesity/overweight and those with normal weight.

Emotional eating has been shown to be associated with multiple psychological symptoms, such as depression, anxiety, and stress (Al-Musharaf, 2020). In the present study, we found positive associations between psychological symptoms and unhealthy eating habits, such as eating in front of TV, replacing main meals with snacks, and snacking, which are habits closely related to emotional eating. Conversely, psychological symptoms were negatively associated with healthier eating habits, such as eating at table. Interestingly, despite emotional eating being more prevalent amongst overweight/obese individuals (Dressler & Smith, 2013), our data suggest that the effects of pandemic on emotional health were comparable between different nutritional statuses, indicating the indistinct need for nutritional support during this period, regardless of the individual’s body weight.

Our study is strengthened by the fact that data was collected during the most severe period of the pandemic in Brazil, thus under the most restrictive stay-at-home orders in place. In addition, this is the first study, to our knowledge, to examine the influence of nutritional status on potential modification in eating habits and food choice determinants. However, this study is not without limitation. The use of self-reported questionnaire may imply some degree of imprecision on data reporting. Also, our sample was predominantly composed by women with high university degree, which limits the generalization of the present findings to less scholarly population.

In conclusion, eating habits were largely modified during the COVID-19 outbreak among Brazilian women, irrespective of nutritional status. Nonetheless, determinants of food choice differed between nutritional statuses. Finally, we showed that determinants of food choice and psychological symptoms were associated with eating habits during the pandemic. The understanding of new eating habits and food choice determinants emerging from the COVID-19 pandemic will help tailor better policies and interventions focused on preventing the risks of food insecurity and overweight in specific populations.

## Data Availability

The datasets used and/or analyzed during the current study are available from the corresponding author on reasonable request.

## Acknowledgements

B.C.M., F.I.S., G.P.E., A.J.P., and B.G. were supported by São Paulo Research Foundation – FAPESP (grants # 2019/14820-6, # 2019/14819-8, #2020/07860-9, #2015/26937-4, and #2017/13552-2). C.F.N. was supported by National Council for Scientific and Technological Development – CNPq (grant #402123/2020-4).

---

**Supplementary Table 1.**
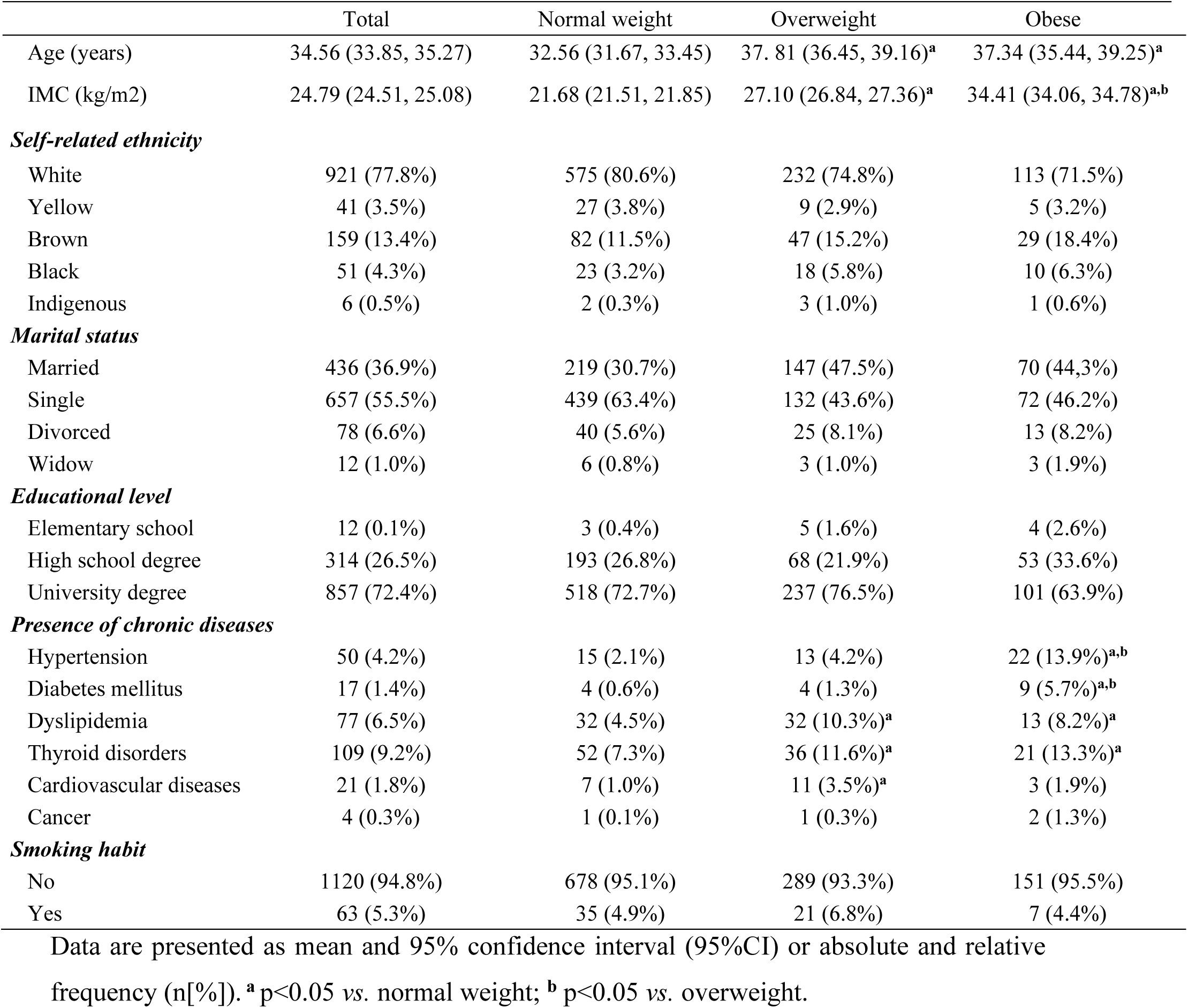
Participants’ characteristics.

|  | Total | Normal weight | Overweight | Obese |
| --- | --- | --- | --- | --- |
| Age (years) | 34.56 (33.85, 35.27) | 32.56 (31.67, 33.45) | 37.81 (36.45, 39.16)<sup>a</sup> | 37.34 (35.44, 39.25)<sup>a</sup> |
| IMC (kg/m<sup>2</sup>) | 24.79 (24.51, 25.08) | 21.68 (21.51, 21.85) | 27.10 (26.84, 27.36)<sup>a</sup> | 34.41 (34.06, 34.78)<sup>a,b</sup> |
| ***Self-related ethnicity*** |  |  |  |  |
| White | 921 (77.8%) | 575 (80.6%) | 232 (74.8%) | 113 (71.5%) |
| Yellow | 41 (3.5%) | 27 (3.8%) | 9 (2.9%) | 5 (3.2%) |
| Brown | 159 (13.4%) | 82 (11.5%) | 47 (15.2%) | 29 (18.4%) |
| Black | 51 (4.3%) | 23 (3.2%) | 18 (5.8%) | 10 (6.3%) |
| Indigenous | 6 (0.5%) | 2 (0.3%) | 3 (1.0%) | 1 (0.6%) |
| ***Marital status*** |  |  |  |  |
| Married | 436 (36.9%) | 219 (30.7%) | 147 (47.5%) | 70 (44.3%) |
| Single | 657 (55.5%) | 439 (63.4%) | 132 (43.6%) | 72 (46.2%) |
| Divorced | 78 (6.6%) | 40 (5.6%) | 25 (8.1%) | 13 (8.2%) |
| Widow | 12 (1.0%) | 6 (0.8%) | 3 (1.0%) | 3 (1.9%) |
| ***Educational level*** |  |  |  |  |
| Elementary school | 12 (0.1%) | 3 (0.4%) | 5 (1.6%) | 4 (2.6%) |
| High school degree | 314 (26.5%) | 193 (26.8%) | 68 (21.9%) | 53 (33.6%) |
| University degree | 857 (72.4%) | 518 (72.7%) | 237 (76.5%) | 101 (63.9%) |
| ***Presence of chronic diseases*** |  |  |  |  |
| Hypertension | 50 (4.2%) | 15 (2.1%) | 13 (4.2%) | 22 (13.9%)<sup>a,b</sup> |
| Diabetes mellitus | 17 (1.4%) | 4 (0.6%) | 4 (1.3%) | 9 (5.7%)<sup>a,b</sup> |
| Dyslipidemia | 77 (6.5%) | 32 (4.5%) | 32 (10.3%)<sup>a</sup> | 13 (8.2%)<sup>a</sup> |
| Thyroid disorders | 109 (9.2%) | 52 (7.3%) | 36 (11.6%)<sup>a</sup> | 21 (13.3%)<sup>a</sup> |
| Cardiovascular diseases | 21 (1.8%) | 7 (1.0%) | 11 (3.5%)<sup>a</sup> | 3 (1.9%) |
| Cancer | 4 (0.3%) | 1 (0.1%) | 1 (0.3%) | 2 (1.3%) |
| ***Smoking habit*** |  |  |  |  |
| No | 1120 (94.8%) | 678 (95.1%) | 289 (93.3%) | 151 (95.5%) |
| Yes | 63 (5.3%) | 35 (4.9%) | 21 (6.8%) | 7 (4.4%) |

Data are presented as mean and 95% confidence interval (95%CI) or absolute and relative frequency (n[%]). <sup>a</sup> p<0.05 vs. normal weight; <sup>b</sup> p<0.05 vs. overweight.

